# A Health Navigator intervention to address the unmet social needs of caregivers of hospitalised children in South Australia: protocol for a mixed-methods pilot study

**DOI:** 10.1101/2025.03.24.25324580

**Authors:** Kate Emily Neadley, Maeve Downes, Lily Chan, Brianna Poirier, John Lynch, Mark Boyd, Cheryl Shoubridge

**Affiliations:** School of Biomedicine, Adelaide University, South Australia; Northern Adelaide Local Health Network, South Australia; School of Medicine, Adelaide University, South Australia; School of Dentistry, Adelaide University, South Australia; School of Public Health, Adelaide University, South Australia

**Keywords:** ‘social determinants of health’, ‘social prescribing’, ‘hospital’, ‘paediatrics’, ‘screening and referral’

## Abstract

**Background:** Interventions that identify patients’ unmet social needs and provide referrals to government and community resources are growing rapidly across healthcare settings. In Australia, research is limited. This protocol paper presents the methodology for a pilot study assessing the feasibility and acceptability of integrating a Health Navigator (HN) intervention into an inpatient paediatric setting that serves a socially disadvantaged population in South Australia.

**Methods:** We will conduct a mixed-methods feasibility and acceptability study of a HN intervention designed to respond to the unmet social needs identified by caregivers of children admitted to the Children’s Ward of the Lyell McEwin Hospital. We will recruit a maximum of 60 participants over a three-month recruitment period, to be assisted by two HNs over a four-month follow-up period. Our primary feasibility outcomes are rates of 1) intervention recruitment, 2) intervention retention and 3) intervention completion. We have selected a ≥80% threshold for feasibility success across all outcomes. Feasibility and acceptability will also be assessed using focus groups with clinicians, participants and community service providers, to explore barriers and enablers to integrating the HN intervention in the ward setting. Secondary outcomes include screening rates for unmet social needs, changes in participants’ unmet social needs pre/post-HN intervention and participant satisfaction with the HN service.

**Discussion:** This study will contribute to the understanding of how HN interventions establish holistic ‘community-facing’ hospitals, connecting vulnerable populations to appropriate resources for the enhancement of participants’ overall health and wellbeing. Our feasibility and acceptability data will inform the design of future trials, as we refine the HN intervention to better suit the needs of socially disadvantaged populations in South Australia.

**Protocol version:** Version 1, date 23/01/2024

**Trial registration:** This trial was prospectively registered in the Australian New Zealand Clinical Trial Registry (registration ID: ACTRN12624000494538).

## Background

It is well-established that experiencing social disadvantage, such as poverty, housing and financial insecurity, can negatively impact parents and caregivers’ abilities to provide secure and nurturing environments in which to parent children (1, 2). This may lead to long-lasting effects on children, as social disadvantage is linked to poorer health and developmental outcomes and educational attainment, and fewer opportunities to develop human and social capital (3, 4). Following the World Health Organisation’s landmark Commission on Social Determinants of Health (4), the role of healthcare settings in mitigating social disadvantage has come under scrutiny. Policies integrating health and social care in countries such as the United States and United Kingdom have resulted in a proliferation of healthcare-based social care interventions, referred to as ‘social prescribing’ or ‘Health Navigator (HN) interventions’ (hereafter referred to as ‘HN interventions’) (5, 6).

### Health Navigator Interventions

HNs are generally non-clinical workers with knowledge of case management principles and the community services sector. Their primary responsibilities are to listen and respond to participants in a non-judgemental and trauma-informed manner as they assist them to connect with appropriate services (5, 6).

HN interventions can be classified as complex interventions, as multiple factors at the individual-, organisational- and community-level impact intervention outcomes (7). Generally, HN interventions comprise three components: 1) participants are screened for unmet social needs, 2) participants reporting unmet social needs are referred to an HN to prioritise needs and create an action plan, and 3) a HN provides participants with referrals and follow-up to appropriate government and community resources (6).

Evidence for HN interventions is mixed: some interventions have been shown to decrease unmet social needs and medication costs, and improve quality of life (8), while others report that patients have little desire to take part in screening or receive referrals to services (9, 10). These mixed outcomes may be due to population characteristics, such as disease type, age, or previous experiences of discrimination in the healthcare system. Differences in intervention settings, e.g. inpatient vs community locations may also contribute (11). HN interventions occur most frequently in primary care, despite the substantial barriers faced by disadvantaged populations to accessing this care (5, 12).

Although Australians benefit from universal healthcare, extensive funding cuts to primary care and the ongoing cost-of-living crisis has limited vulnerable children and families’ access to preventive care, creating increased demand on hospital and emergency services (13, 14). Hospitals provide an important opportunity to engage with vulnerable children and caregivers and HN interventions targeting caregivers provide a unique opportunity for early intervention in child health and wellbeing. In Australia, HN intervention research is nascent, with most studies exploring screening for unmet social needs with limited referral pathways (15, 16). Here we present a mixed-methods study protocol to assess the feasibility and acceptability of embedding an HN intervention in an inpatient paediatric setting to address caregivers’ social needs.

## Methods

### Primary Aim

To explore the feasibility and acceptability of a HN-led intervention to identify and address the unmet social needs of caregivers of hospitalised children.

### Secondary Aims

1) To co-develop communication guidelines with community and clinicians to assist with screening for unmet social needs in the ward
2) To explore patterns of screening for unmet social needs in the presence and absence of the HN in the ward
3) To explore changes in the prevalence and type of unmet social needs pre- and post-HN intervention
4) To explore total number and type of services accessed by participants pre- and post-HN intervention

### Setting

The intervention will be conducted in the Children’s Ward of a major hospital that serves a disadvantaged population in Adelaide, South Australia (17). Many caregivers of children admitted to this service are single parents and low-income earners (17), and approximately 20% of children have contact with the Child Protection (CP) system before they reach school(18). Healthcare settings are one of the largest sources of CP notifications (19), which can cause caregivers to distrust clinicians as they fear CP involvement. Despite this social complexity, our previous research suggests that caregivers of children admitted to the Children’s Ward have an overall positive view of screening for unmet social needs and are willing to disclose social risks (20).

### Design

Following the latest Medical Research Council guidelines for complex interventions (7), we selected a. exploratory, mixed-methods feasibility study design, utilising pre–post intervention assessments and qualitative and quantitative assessments of participant and stakeholder groups.

This will involve:

1) Engagement with caregivers and consultation with ward team members to inform HN service integration in the ward
2) Pre-post intervention audits of screening rates for unmet social needs
3) Pre-post intervention assessments of clinician feasibility and acceptability measures
4) Post-HN intervention interviews with ward team members, community service providers and participants to explore experiences with the HN intervention.

Inclusion Criteria:

- Parent(s)/caregiver(s) of children (0 to <18 years) admitted to the Children’s Ward in the LMH

Exclusion Criteria:

- Parent(s)/caregiver(s) of children readmitted to the ward that have previously been recruited to the HN intervention
- Parent(s)/caregiver(s) <18 years
- Limited English proficiency, as research funds are insufficient to hire interpreters
- Parent(s)/caregivers actively involved with the Child Protection Services or intensive family support services
- Children placed in out of home care

The study design adheres to Standard Protocol Items: Recommendations for Interventional Trials (SPIRIT) guidelines (21), including a SPIRIT schedule of intervention activities (Table 1) and SPIRIT checklist (see Additional File 1).

**Table 1:**
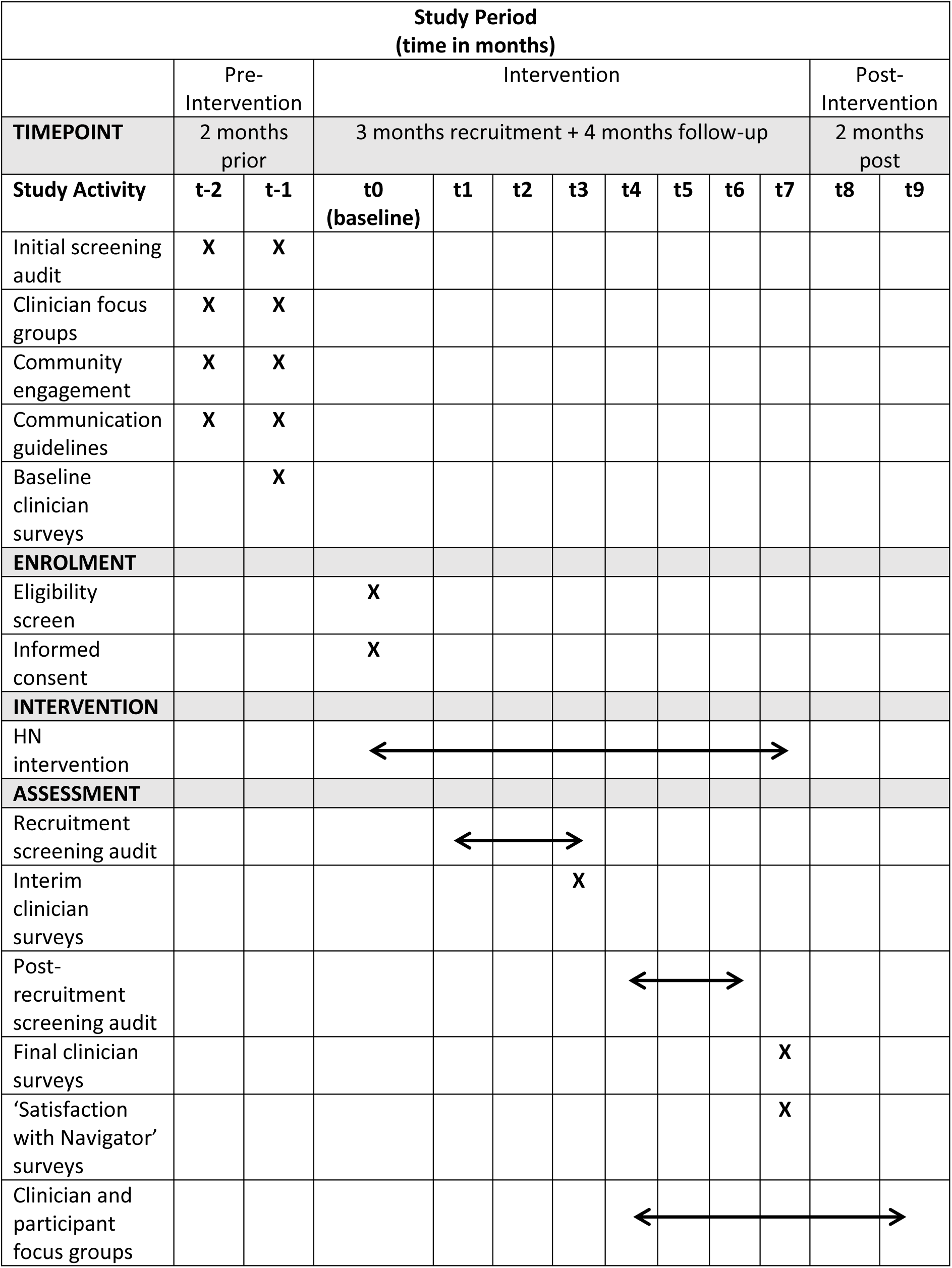
SPIRIT schedule of enrolment, intervention activities and assessments.

### Community & clinician engagement

Clinicians screen caregivers for unmet social needs as standard practice during the admission process. This screening uses a refined version of the Unmet Needs Screening Tool (22) co-designed with clinicians to best meet the needs of families presenting to the ward. Ward team members report discomfort conducting screening as they feel they lack appropriate training to ask sensitive social needs questions and are wary of damaging relationships with caregivers (23, 24). To further explore clinicians’ views of screening, prior to the HN intervention, ward team members will be invited to take part in focus groups concerning their experiences of and perceived barriers to screening for unmet social needs. As caregivers admitted to the ward are unlikely to be able to attend focus groups due to caring responsibilities, a researcher (KN) will facilitate one-on-one semi-structured interviews with caregivers to explore possible improvements to the screening process. Both caregivers’ and ward team members’ suggestions will be compiled into a set of brief communication guidelines attached to the screening tool. These guidelines will assist clinicians to ask sensitive social needs questions.

### Feasibility & Acceptability: Children’s Ward team surveys

Children’s ward team members include clinicians, play coordinators and education members. Evidence from paediatric settings suggests that ward team members are more likely to screen caregivers of children admitted to the ward for their unmet social needs when there is a dedicated team member than can provide participants with support for their reported needs (24, 25). To better understand how ward team members feel about conducting screening and referral for unmet social needs in the ward, we will administer online, anonymous surveys containing brief feasibility and acceptability measures (FAIMs) (15) (see Additional File 2). FAIMs assess five key facets of intervention feasibility and acceptability, e.g. if ward team members believe screening is important or easy to complete, using a three-point Likert scale: ‘Disagree’, ‘In the Middle’ and ‘Agree’. FAIMs will be administered electronically via email to all ward team members pre-intervention, during recruitment and post-recruitment when the HN is removed from the ward. FAIMs take no longer than three minutes to complete and all responses will be anonymous (Table 1).

### The HN Intervention

Ward team members will administer the screening tool to caregivers, guided by caregivers’ preference to either self-complete or receive assistance from a ward team member when completing the tool. For caregivers who have completed screening and request assistance with reported needs, and are eligible for the study, will be recommend by a ward team member to the HN. If the eligible caregiver consents, the HN will meet with the eligible participant and obtain written, informed consent. The HN will obtain written informed consent and conduct an initial meeting with the caregiver to understand their current situation and co-develop a referral plan to appropriate community and/or government resources. If the caregiver requires any assistance making contact, the HN will assist by:

- Making phone calls to community service providers
- Organising meeting times
- Organising transport
- Attending meetings to advocate for the participant

If the caregiver does not wish to have this initial meeting in the Ward, the HN will make contact via telephone within 14 days of recruitment. Follow-up will be scheduled for monthly contact over a four-month follow-up period (see Table 1). This follow-up schedule can be reasonably adjusted by the participant as preferred. At each follow-up contact, the HN will seek information from the caregiver regarding the status of their unmet social need(s) to inform their follow-up procedures (Table 2).

**Table 2:** Status of participants’ unmet social needs and appropriate HN follow-up procedures.

|  |  |  |
| --- | --- | --- |
| **Unmet social need status** | **Definition** | **HN Action** |
| **Resolved** | Social needs have been met. | HN will continue service with any remaining unmet social needs. |
| **Engaged** | Participant requires more time with the resource. | HN will revisit in the next follow-up call. |
| **Failed** | The resource was not useful or unable to meet the participants' needs. | Another resource will be identified, if possible, by the HN to assist the participant. |

Caregivers will be able to call the HN at any time during their work hours by study phone if they require assistance. Caregivers will be classified as ‘lost to follow-up’ if the HN is unable to contact the caregivers after three attempts in one month. These three attempts will be made using different modes of contact: SMS, email, and phone, to allow for any difficulties participants may be experiencing. If caregivers request to cease follow-up before the four-month endpoint because they have been appropriately engaged with services, this will be recorded as ‘early exit’.

At the end of HN follow-up, caregivers will be asked to complete unmet social needs screening. HNs will facilitate personalised referrals (‘warm handovers’) to partners in the community for unresolved or emerging unmet social needs, ensuring they receive ongoing support. Post-intervention, a researcher will administer via SMS a brief ‘Participant Satisfaction with Health Navigator’ (PSN) questionnaire, modified from existing measures (26), to explore caregivers’ satisfaction with the HN service (Additional File 3). PSN measures assess nine important aspects of HN performance, e.g. the development of trust and respect with participants, and quality of communication. As recommended by ward team members, we tailored PSNs to the population by reducing the original five-point Likert scale to a simplified three-point scale (‘Disagree’, ‘In the Middle’ and ‘Agree’). Both FAIMs and PSNs will be administered electronically, either via SMS or email, to participants using Qualtrics software, version XM (Qualtrics, Provo, UT), supplied by Adelaide University.

### Responsibilities and Limitations of the HN Role

As the ultimate aim of this research is to implement the HN role in the hospital setting, HNs will follow the same mandatory reporting requirements as all ward team members to reflect real-life role implementation. Similar to other hospital-based interventions, the HN will work in conjunction with hospital social workers and the HN role and not provide direct HN support for mental health crisis or family and domestic violence (FDV) (23, 27, 28). Instead, if caregivers experience crisis, both in the hospital and in the community, HNs will follow Standard Operating Procedures to escalate to ward team members and/or study investigators as appropriate.

We expect the HN role will be emotionally challenging at times. For debriefing and assistance with case complexity, HNs will receive supervision by a senior social worker for at least one hour per week.

### Special considerations

We anticipate clinician referrals to the HN may be a barrier to recruitment, as clinicians may not understand the HN role or lack sufficient time to make referrals. A rotating roster of casual staff may cause further difficulties, as new team members are introduced to the ward throughout the intervention period (25). To improve clinician’ awareness of the intervention, HNs will be embedded into the ward multidisciplinary team and take part in team ‘huddles’, facilitating discussions around consenting participants’ unmet social needs as appropriate. A member of the research team will also be present on the ward to contact ongoing education sessions with ward team members to address concerns and co-develop solutions to any issues that may arise.

### Feasibility & Acceptability: Auditing screening and referral processes for unmet social needs

Clinicians report greater likelihood of conducting screening when they are able to provide patients with appropriate assistance for their disclosed unmet social needs (24, 25). Audits of social needs screening rates in the presence and absence of HN support will provide insight into screening acceptability for ward team members, as greater screening completion should indicate screening is acceptable and feasible. We will conduct an audit of screening rates and completeness at the following time points (see Additional File 4 for audit items):

1. **Pre-intervention Audit -** Focusing on the same three-month period in which recruitment is scheduled, we will audit screening records from a previous year to avoid potential contamination of screening rates by current research efforts. Due to complications with the introduction of the electronic medical record in 2023, we will audit records from June-August 2022.
2. **Recruitment Audit –** The HN will be present on the ward throughout the three-month recruitment period (June-August 2024). We will audit screening records for caregivers admitted to the ward during this period.
3. **Post-recruitment Audit –** Once HNs have complete caseloads, they will be removed from the ward. We will audit screening across all patients admitted to the ward when the HN is no longer present assist with referrals, from September-November 2024.

We estimate approximately 1000 admissions over a 3-month period. Assuming a sample screening proportion of 0.5 (the most conservative approach), and a margin of error of 5% at the 95% confidence level, 278 admissions from an estimated total 1000 admissions will be needed. We therefore aim to audit 300 admissions for each 3-month auditing period.

### Analysis

#### Sample size

Following the most recent guidelines for evaluating intervention feasibility, practical considerations, such as participant flow, were used to generate sample size (29, 30). Our sample size will be determined by HN workload. Community-based organisations performing similar roles recommend an HN caseload of no more than three participants per day to allow sufficient time for research and follow-up in the community. Two HNs will service a 1.0FTE role, limiting our sample size to 30 participants per HN, for a maximum sample of 60 participants.

#### Primary Outcome: quantitative analysis of intervention feasibility and acceptability

A mixed-methods approach to analysis is required to best capture the complexity of feasibility and acceptability outcomes (7). We selected process measures, i.e. intervention recruitment, retention and completion, as quantitative measures of intervention feasibility (Table 3). Time points at which process measures will be calculated are detailed in Additional File 5. A threshold of 80% for each measure is considered an appropriate estimate for feasibility success (30, 31).

**Table 3:** Intervention feasibility as defined by process measures, their definitions and criteria for feasibility success.

| Process measure | Definition | Success Criteria |
| --- | --- | --- |
| **Recruitment rate** | Number of caregivers who request assistance with reported unmet social needs and consent to HN intervention, divided by the total number of caregivers who request assistance with reported unmet social needs (with or without consent to the HN intervention) | ≥80 % of eligible participants recruited |
| **Retention rate** | Number of participants who complete follow-up per protocol, i.e. one appointment/follow-up month, divided by the total number of participants who have commenced the HN intervention and complete less or more than the scheduled one appointment/follow-up month | ≥80 % of participants complete four follow-up appointments (one appointment/follow-up month) with the HN |
| **Completion rate** | Total number of participants who complete intervention divided by the total number of participants who have commenced the HN intervention | ≥80 % of participants complete HN intervention |

Ward team members’ and participants’ views of intervention acceptability will be assessed using FAIMs (see Additional File 2) and PSN measures (Additional File 3) respectively. Scores for FAIMs and PSNs (‘Disagree’=0, ‘In the Middle’=1 and ‘Agree’=2 points) will be calculated for each survey item. Total scores for FAIMs will be compared across three study phases: pre-intervention, recruitment and post-recruitment. The intervention will be considered feasible and acceptable for ward team members if 80% of team members ‘agree’ (score=2) that screening is 1) important and 2) possible during the recruitment phase. The HN intervention will be determined as acceptable to participants if PSN scores equal a minimum of 14 for each participant (nine survey items with maximum score= 18, 80%= 14). Simplified three-item Likert scales were selected to minimise the likelihood of missing data, and any missing data will be omitted.

#### Primary Outcome: qualitative analysis

Quantitative analysis alone is insufficient for robust evaluation of a complex intervention (7). We will assess clinicians’, caregivers’ and community service providers’ views of the HN intervention through separate focus groups and/or one-on-one interviews, as appropriate. Interviews and focus groups will follow a semi-structured interview schedule and will be explored using codebook thematic analysis (TA) (32, 33). The data corpus (transcripts from caregivers, ward team members and participants) will be combined and explored using NVivo 12 software (QSR International Pty Ltd. Version 12.6.1). Codebook TA was selected as authors highlight the importance of researcher subjectivity in coding and interpretation of data. Codebook TA will be used to identify latent and semantic themes concerning barriers and enablers to intervention uptake, and participants’ and ward team members’ experiences of the HN intervention. Codebook TA is philosophically incompatible with positivist quality control measures such as intercoder reliability (33). Rather, to ensure rigorous analysis standards are upheld, two researchers (KN and BP) will sequentially code and discuss identified themes . Key themes and relevant data extracts will be presented in tabular form.

Qualitative and quantitative data will be analysed concurrently, and a weaving approach will be used to present barriers and enablers to intervention on a theme-by-theme basis (34).

#### Secondary Outcomes

We expect rates of screening for unmet social needs will improve when the HN is on the ward during the recruitment phase. Screening rates (the number of screening tools completed) and screening completeness (proportion of screening questions completed) will be explored: 1) pre-intervention, 2) during recruitment when the HN is readily available on the ward, and 3) when the HN is conducting follow-up in the community (see section: ‘Auditing screening and referral processes for unmet social needs’). To ensure a random sampling approach, we will assess the first 100 screening tools from each month of the audit period. To explore how HNs impact participants’ unmet social needs, the social needs screening tool will be conducted at baseline and repeated on follow-up completion. The number of social needs engaged with a service, resolved and failed to be resolved by the HN will be explored at follow-up end. HN impact on connecting participants to appropriate services will be assessed by comparing the total number and type of services accessed by caregivers at the time of the Initial Meeting and follow-up end (Table 4).

**Table 4:**
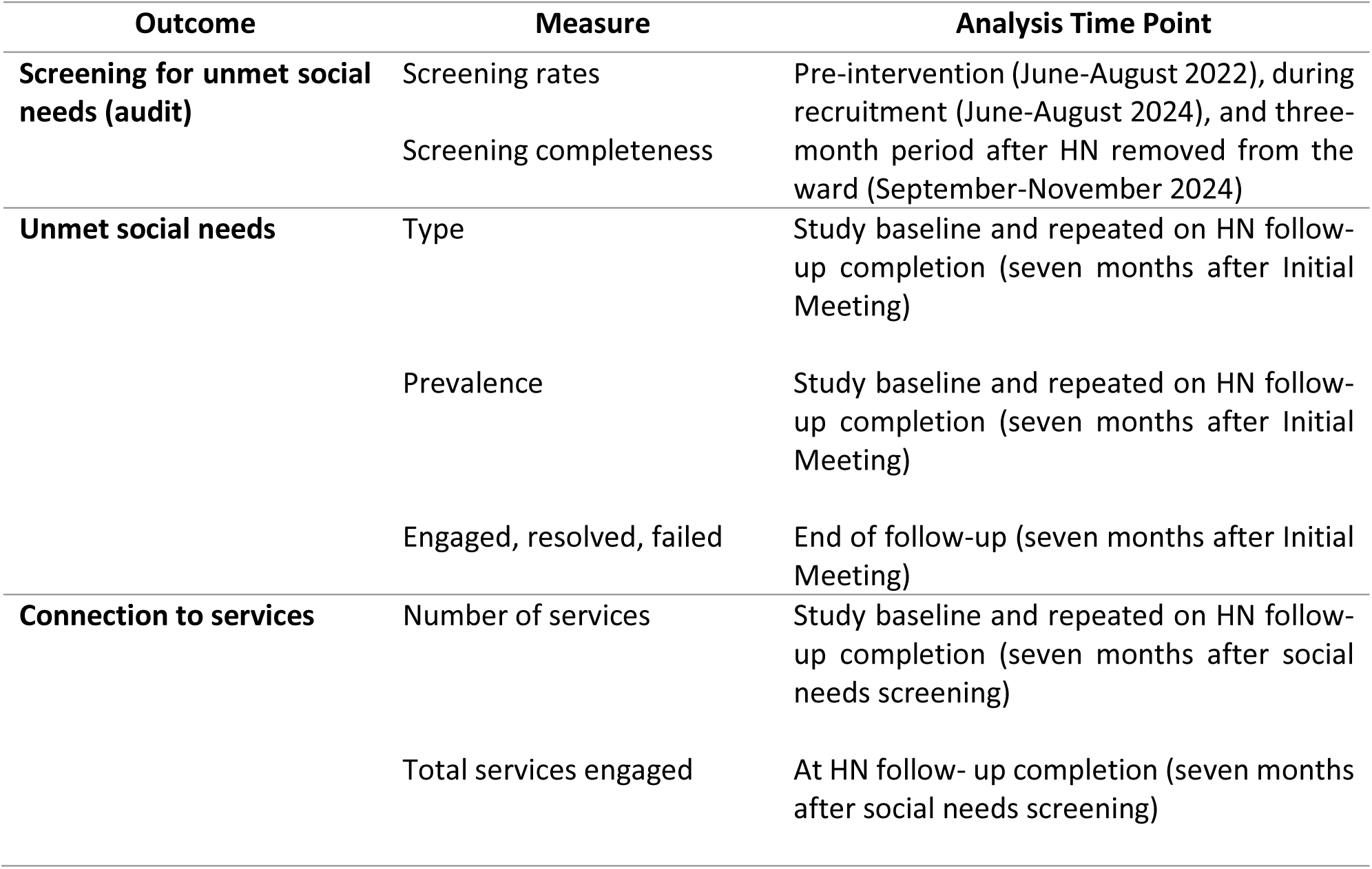
Secondary outcome measures and analysis time points during the HN intervention.

### Ethical Considerations

#### Potential benefits and risks

Potential benefits to participants include assistance with unmet social needs, e.g. assistance and advocacy accessing community services to ameliorate housing and financial insecurity. These benefits may extend to the local community, as the study will identify referral pathways for unmet social needs that can be captured and used by clinicians and other members of the community to access assistance with unmet social needs. Potential risks to participants may include experiencing emotional distress discussing sensitive unmet social needs (e.g. FDV, homelessness) or during follow-up whilst accessing community resources. HNs will caution participants that we may be unable to provide assistance due to a lack of available community resources. Caregivers on the ward who disclose social needs but decline to participate in the study, or those who disclose needs after the intervention has ceased on the ward, will be provided a referral to the social work department, as would occur during standard care.

### Confidentiality and access to data

Participants’ identifying data will be stored in hard copy and on a password-protected document that links their information and study identifier. Only investigators will have access to this identifying document. All intervention and follow-up data will be de-identified and stored using Research Electronic Data Capture (REDCap) software hosted at The University of Adelaide software. REDCap is a secure, web-based software platform designed to support data capture for research studies, providing 1) an intuitive interface for validated data capture; 2) audit trails for tracking data manipulation and export procedures; 3) automated export procedures for seamless data downloads to common statistical packages; and 4) procedures for data integration and interoperability with external sources (35).

We do not anticipate this intervention will result in serious adverse events, therefore a Data Monitoring Committee will not be required.

## Discussion

To our knowledge, this protocol paper presents the methodology for the first inpatient paediatric HN intervention in Australia. Our research will provide valuable data on current priorities in the field, including how to integrate HN interventions in a hospital setting, elucidating community referral pathways for unmet social needs and contributing to the understanding of HN roles and limitations (6, 36). Our research is limited by the lack of non-English speaking participants, but the feasibility and acceptability data we collect will inform the design of larger, more inclusive trials of HN interventions in paediatric settings if the intervention is appropriate to ward team members and participants (29). With community in mind, this research will deepen understanding of the unmet needs and social disadvantage experienced by families of children in this disadvantaged area of South Australia. This information could in turn be used to advocate for of targeted support services. We will build relationships with community service providers and create better hospital-community linkages, taking first steps towards a ‘community-facing’ hospital providing holistic support for patients’ health and wellbeing.

## Supporting information

Supplementary File 1 SPIRIT checklist

## Data Availability

All data produced in the present study are available upon reasonable request to the authors

## List of Abbreviations

HN: Health Navigator
CP: Child Protection
FAIM: Feasibility and Acceptability Intervention Measures
FDV: Family and domestic violence
PSN: Participant Satisfaction with Health Navigator measure

## Declarations

### Ethics approval and consent to participate

This research received ethical approval from the Central Adelaide Local Health Network Human Research Ethics Committee (approval no: 18968). All participants will provide informed, written consent.

### Consent for publication

All participants will provide informed, written consent to participate in this research. Only de- identified data will be published. We will publish results in open access, peer-reviewed scientific journals and disseminate findings to clinicians and participants upon completion of analysis.

### Availability of data and materials

The datasets used and/or analysed during the current study are available from the corresponding author on reasonable request.

### Authors’ contributions

All authors are responsible for trial conceptualisation and design. LC provided analysis and biostatistics support. All authors read and approved the final manuscript.

## Acknowledgements

We would like to acknowledge the ongoing support and advocacy of the Children’s Ward team. Our research would not be possible without your drive to go above and beyond in assisting these vulnerable children and caregivers.

