## Supplementary files 2, 3 and 5 for "A Health Navigator intervention to address the unmet social needs of caregivers of hospitalised children in South Australia: protocol for a mixed-methods pilot study"

### Additional File 2: Feasibility and Acceptability Intervention Measures

**Feasibility Intervention Measure**

1. Screening and referral for unmet social needs seems possible

Disagree	Neutral	Agree

2. The screening tool for unmet social needs is easy to complete

Disagree	Neutral	Agree

3. Providing referrals for assistance for unmet social needs is easy

Disagree	Neutral	Agree

**Acceptability Intervention Measure**

1. I think screening and referral for unmet social needs is important

Disagree	Neutral	Agree

2. I support screening and referral for unmet social needs in the Children’s Ward

Disagree	Neutral	Agree

### Additional File 3: Participant Satisfaction with Health Navigator (PSN)

1. My Health Navigator gives me enough time

Disagree	In the Middle	Agree

2. I trust my Health Navigator

Disagree	In the Middle	Agree

3. I can rely on my Health Navigator

Disagree	In the Middle	Agree

4. My Health Navigator respects me

Disagree	In the Middle	Agree

5. My Health Navigator listens to me

Disagree	In the Middle	Agree

6. My Health Navigator is easy to talk to

Disagree	In the Middle	Agree

7. I feel like my Health Navigator cares about me

Disagree	In the Middle	Agree

8. My Health Navigator helps me figure out answers to my problems

Disagree	In the Middle	Agree

9. My Health Navigator is easy for me to reach

Disagree	In the Middle	Agree

### Additional File 5: Participant Flow Diagram and Process Measures

**Figure 1. Participation flow diagram and process measures.** Colours highlight study time points at which process measures are calculated. Pink refers to recruitment rate, orange for rate of intervention uptake, and green for intervention completion. The dark green colour refers to intervention retention, i.e. the number of participants who complete more or less than the predicted four follow-ups (one appointment/month of follow-up) with the HN.

HN: Health Navigator

Carers completed screening tool n= number of admissions during recruitment period

Carers with no unmet needs n=

Carers with unmet needs, requested assistance n=

Carers not referred to HN by clinicians n=

Reasons: Did not meet study criteria, n= Reached capacity, n=

Carers referred to HN by clinicians n=

Did not meet study criteria n=

Met study criteria n=

Not approached by HN n=

Reasons: Reached capacity, n= Not contactable, n=

Approached by HN n=

Did not consent to study n=

Reasons: Not interested, n= Not eligible, n=

Consented to study n=

No initial or subsequent meeting n=

Reasons: Not contactable, n= Withdrew, n=

Commenced intervention n=

Did not complete intervention n=

Reasons: Not contactable, n= No longer eligible, n= Withdrew, n=

Completed intervention including early exit n=

Did not complete Satisfaction with HN measures n=

Reasons: Declined, n= Not contactable, n=

Completed Satisfaction with HN measures n=

Not satisfied with HN n=

Satisfied with HN n=

Carers with unmet needs, did not request assistance n=
